# Determinants of Anemia among school children and adolescents in Zanzibar

**DOI:** 10.1101/2025.02.28.25323108

**Authors:** Patrick Codjia, Germana Leyna, Hawa Msola, Gibson Kagaruki, Geofrey Mchau, Tedson Lukindo, Ray Masumo, Erick Killel, Asha Salmin, Fatma Ally Said, Adam Haji Ali, Kombo Mdachi Kombo, Anna Mosses, David Solomon, Esther Ngadaya, Joyce Ngegba

## Abstract

**Background:** Anemia continues to pose a substantial global health concern. Previous research in Tanzania mainland has documented high anaemia prevalence among children. However, studies specifically exploring the state of anaemia among school-aged children in Zanzibar remain limited.

**Objective:** To determine the prevalence and determinants of anaemia among children and adolescents aged 5 to 19 years in Zanzibar.

**Methods:** The Zanzibar National School Health and Nutrition Survey was aschool-based cross-sectional survey among school children and adolescents aged between 5 and 19 years enrolled in primary and secondary schools during the academic year 2022. A multistage systematic sampling was applied to select primary schools, class levels and students. Pre-tested structured questionnaires were used to gather demographic and economic data. Hemoglobin concentration was determined using a hemoglobinometer (HemoCue 301+). Inferencial analysis was done using Chi-Squire test and Modified Poison Logistic Regression.

**Results:** Almost half of school children and adolescents 5-19 years in Zanzibar are anaemic with a prevalence of 45.7%. The determinants of anaemia included gender, age, school ownership, meal frequency and family income. Children aged 5-9 years were 1.2 more likely to be anaemic (APR: 1.2, 95% CI: 1.1-1.4; p<0.001). Adolescent girls had 10% times more significant risk of anaemia than adolescent males (APR: 1.1, 95% CI: 1.1-1.3; p=0.001). Children in public schools had a 1.7 times significantly higher risk of being anaemic than their counterparts in private schools (APR: 1.7, 95% CI: 1.2-2.3; p=0.004). Children who consumed breakfast 2-4 times a week had a significantly decreased risk of anaemia (APR: 0.8, 95% CI: 0.7-0.9p=0.015). Wealth quintile, settings, School level, regional differences, urban and rural settings, and deworming status, showed no significant difference in anaemia prevalence.

**Conclusion:** Despite massive improvements as compared to previous studies, the findings indicate a persistently higher prevalence of anaemia among school children and adolescents in Zanzibar. Both school-and-community focused interventions are needed to address the burden of anaemia in Zanzibar. The preponderance of anaemia among adolescent girls and those from public schools provides a unique opportunity for targeting such populations with anaemia prevention and control interventions, and schools are uniquely positioned to provide such interventions.

## BACKGROUND

Anaemia, characterized by a decrease in the number or size of red blood cells continues to pose a substantial global health concern[1]. It has been estimated to affect 24.8% of the world’s population, with a particularly high prevalence in low-income countries[2]. Among children, the statistics are even more alarming. According to the World Health Organization (WHO), nearly 43% of children under five years of age suffer from anaemia, and in sub-Saharan Africa, this figure increases to a staggering 63% in pre-school children, and 48% among school-aged children[3, 4].

The aetiology of anaemia is multifactorial and often includes nutritional deficiencies, infectious diseases, and genetic disorders[5]. Among nutritional deficiencies, iron deficiency is the most common cause, but deficiencies in vitamins such as B12 and folic acid also play significant roles[6]. Infectious diseases, including malaria and soil-transmitted helminth infections, represent substantial contributors to the anaemia burden, especially in tropical and subtropical regions [7, 8]. Furthermore, socio-economic and demographic factors, including low maternal education, poor dietary diversity, low household income, and limited access to healthcare services, have also been strongly associated with the prevalence of anaemia in children[9]. Research has shown that anaemic children often have a poorer cognitive and physical development trajectory, leading to adverse outcomes on their educational attainment and future economic productivity [10].

In the United Republic of Tanzania, the burden of anaemia has been a long-standing public health issue. Prior research in mainland Tanzania has documented high anaemia prevalence among children, reaching up to 72% in some regions[11]. However, studies specifically exploring the state of anaemia among school-aged children in Zanzibar remain limited.

Identifying age-stratified profiles of anaemia could provide key insights into the life-stage-specific prevalence and intensity of anaemia. Furthermore, pinpointing the contributing factors can aid in directing effective and targeted health policies and interventions for anaemia prevention and control, specifically in susceptible populations such as school-going children. Our study aims to fill this knowledge gap by investigating the age-stratified profiles and determinants of anaemia among children and adolescents aged 5 to 19 years in Zanzibar.

## METHODS

### Study design

The Zanzibar National School Health and Nutrition Survey (SHNS) was a school-based cross-sectional survey among students aged between 5 and 19 years who were enrolled in primary and secondary schools during the academic year 2022(1).

### Study setting

The study was conducted in all the eleven administrative districts of Zanzibar, with a total population of 1,889,773 persons according to the 2022 Census [12]. Over one-third of the population is between 5 to 19 years of age. Zanzibar’s marine ecosystem is an important part of the economy for fishing and algaculture and contains important marine ecosystems that act as fish nurseries for Indian Ocean fish populations, and also the main source of protein for its citizens including children and adolescents.

### Sampling technique and sample size

We utilized a stratified multi-stage sampling design [13]. The initial stage involved selecting a sample of schools as the primary sampling unit (PSU) using a probability proportional to size (PPS) method. In the second stage, one class level was selected using a simple random sampling approach (SRS), followed by the selection of individuals from these classes. Each selected class had 28 students evenly distributed by sex (14 girls and 14 boys), chosen using a systematic sampling technique. The sample size was estimated at the district level (domains) and aggregated to obtain the regional and national samples.

### Data and Sample collection

#### Social-demographic and economic data collection

A pre-tested structured questionnaire was used to collect data on demographic characteristics and socioeconomic factors after obtaining written informed consent form through face-to-face interview. Questionnaire were developed in English, translated into Swahili and then back-translated to English to verify accuracy. Independent variables such as household wealth index was assessed as an indicator of socio- economic status according to a standard approach in equity analysis [21]. Durable household assets indicative of wealth (i.e., telephone, radio, TV, refrigerator, lantern, cupboard, houses with electricity, motorcycle, bicycle, cart etc.) were recorded as (1) “available and in working condition” or (0) “not available and/or not in working condition.” These assets were analysed using principal components analysis (PCA). The component resulting from this analysis was used to categorize households into two approximate quintiles with the following categories: Lowest, Second, Middle, Fourth and Highest quintile

#### Haemoglobin determination

Sterile needles were used to collect less than 300µl of blood from each pupil’s finger-prick. Hemoglobin concentration was determined using a drop of blood stored in microcuvettes, which were inserted into a hemoglobinometer (HemoCue 301+, Angelholm, Sweden) which is recommended by WHO for the use of field surveys [14]. Two trained senior laboratory technologists were responsible for taking specimens for laboratory testing. Haemoglobin (Hb) levels were adjusted for altitude in survey enumeration areas > 1,000 metres above sea level and measured in grams per decilitre (g/dl). Anaemia levels were classified according to the age and sex of adolescents. Table 1 below-defined haemoglobin levels used to diagnose anaemia among the study participants.

**Table 1:**
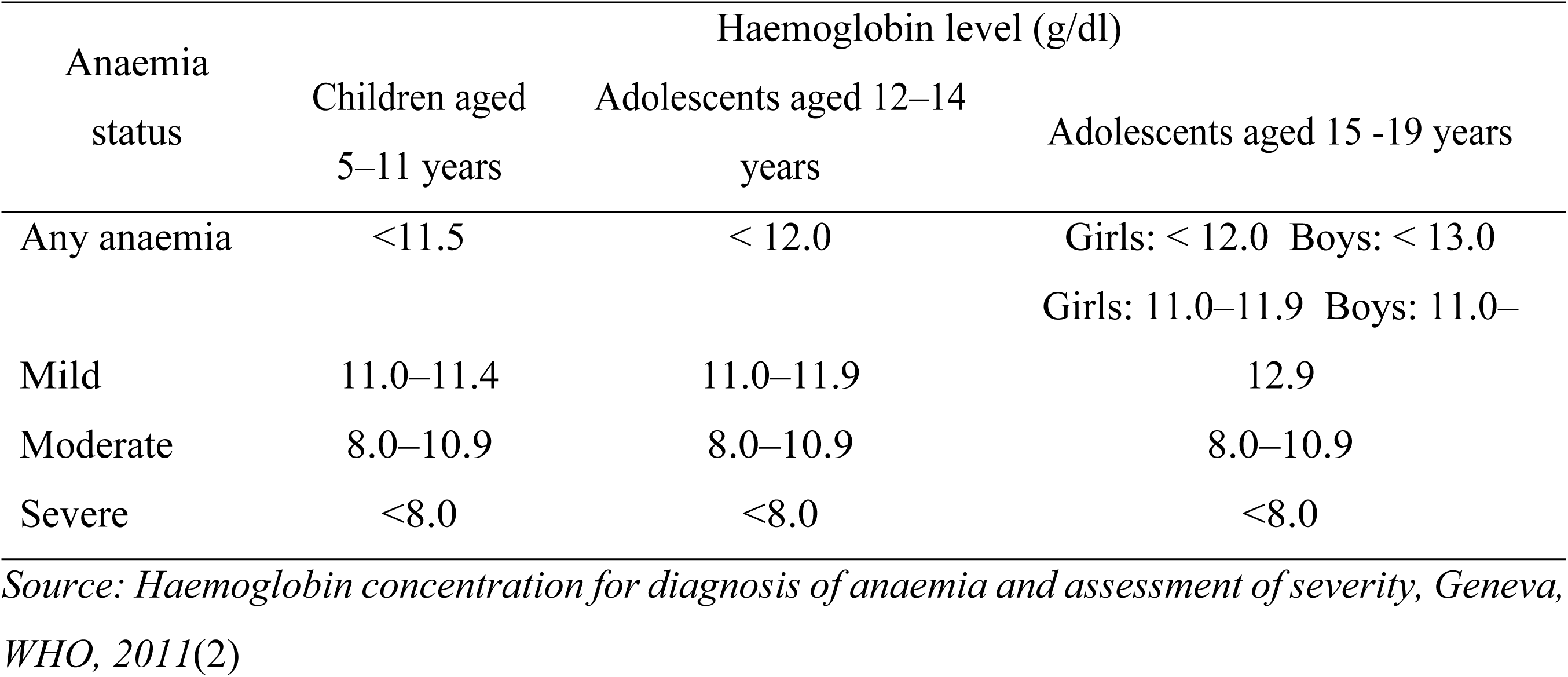
Haemoglobin levels to diagnose anaemia among children and adolescents aged 5 to 19 years.

### Examination of other diseases

#### The Prime Diet Quality Score (PDQS)

Dietary quality as an optimal measure of nutrients for the well-being of schoolchildren and adolescents 5-19 years was assessed using the Prime Diet Quality Score (PDQS). The PDQS was recently developed using a modified Prime Screen questionnaire as a way to characterize diet quality globally [15]. PDQS contained 21 food groups; 13 were categorized into healthy, and 8 as unhealthy food groups. Food groups were queried on a 0- 5 point Likert scale: 0= never, 1= once in a week, 3=2-4 times a week, 4=5-6 times a week and 5=every day. Points were assigned for consumption of healthy food groups as follows: 0–1 serving/week 0 points; 2–3 servings/week 1 point; and ≥4 servings/week=2 points. Scoring for unhealthy food groups was assigned as follows: 0–1 serving/week=2 points; 2–3 servings/week= 1 point; and ≥4 servings/week= 0 points. Individual scores were aggregated to generate an overall score which ranged from 0 to 42 possible scores, the higher the score the higher the quality of diet consumed by a respective children or adolescent.

#### Dietary Habits

Dietary habits among school children aged 5-9 and adolescents aged 10-19 years were assessed using two items, having breakfast and eating snacks seven days before the survey.

### Data management and analysis

#### Data management and analysis

Quantitative data was collected using tablets with installed ODK programmed software. A chain of supervisors approved the data, transmitted to the cloud server after every session of data entry. The data was downloaded and exported into Stata version 17 (STATA Corporation, TX-USA) for cross-checking, data cleaning and analysis. De-identification of personal information was done to ensure anonymity and confidentiality. Data cleaning was undertaken by a trained team. The prevalence of anaemia and the nutritional status of school children and adolescents were interpreted based on anthropometric measurements using specific cut-off points for each age group category [1]. Data analysis started with running descriptive statistics to describe the characteristics of the data set. This included univariate analysis on frequency and percentage response distributions, as well as measures of central tendency and dispersion. Later we conducted inferential analysis using appropriate methods (e.g., Chi-Square test and Modified Poison Logistic Regression). We conducted bivariate analysis to assess a difference in proportion outcomes e.g. prevalence of anemia among school going children. Modified Poison Logistic regression is recommended when fitting outcome with proportion ≥ 10%. Therefore, logistic regression is not suitable as it overestimates odd ratios when the outcome of interest is ≥10% (Zou; 2004 & Lee, 2009). With Modified Poison, we will report both unadjusted and adjusted prevalence ratio (PR). We computed the crude and adjusted estimates. All explanatory variables with a p-value <0.2 in the crude analysis were included in the multivariable model. However, only variables with a p-value <0.05 were retained in the final model and considered to be statistically significant.

#### Ethical Considerations

Ethical clearance for the study was obtained from the Zanzibar Medical Research and Ethics Committee. Special care was taken to protect vulnerable participants (under 18), and consent was secured from schools and parents/guardians. The study team was trained in human subject protection. Consent and assent were sought from the target group of 5–19-year-olds. There were minimal risks associated with participation, with the main concern around discomfort during phlebotomy.

## RESULTS

### Demographic characteristics

The study was implemented in all 11 districts of Zanzibar. The demographic and socio-economic characteristics of the study participants are presented in Table 2 . A Slightly higher proportion (51.7%) of the study participants were girls. Almost 2 in 5 were individuals from 10-14 years, and majority (69%) were from primary school and reside in urban areas.

**Table 2:** Demographic and socio-economic characteristics of the study participants (N=2,556).

| Characteristics | Number | Percentage (%) |
| --- | --- | --- |
| **Sex** |  |  |
| Male | 1,234 | 48.3 |
| Female | 1,322 | 51.7 |
| **Age (years)** |  |  |
| 5 – 9 | 784 | 30.7 |
| 10 – 14 | 1,058 | 41.4 |
| 15 – 19 | 714 | 27.9 |
| **Level of Education** |  |  |
| Primary | 1,768 | 69.2 |
| Secondary | 788 | 30.8 |
| **School type** |  |  |
| Public | 2463 | 96.4 |
| Private | 93 | 3.6 |
| **Area** |  |  |
| Rural | 1,194 | 46.7 |
| Urban | 1,362 | 53.3 |
| **Wealth quintile** |  |  |
| Lowest | 530 | 20.7 |
| Second | 493 | 19.3 |
| Middle | 513 | 20.1 |
| Fourth | 538 | 21.0 |
| Highest | 482 | 18.9 |
| **District/Region** |  |  |
| North A district | 184 | 7.2 |
| North B district | 196 | 7.7 |
| **Unguja North** | **380** | **14.9** |
| Central district | 196 | 7.7 |
| South district | 196 | 7.7 |
| **Unguja South** | **392** | **15.3** |
| Urban district | 264 | 10.3 |
| West A district | 305 | 11.9 |
| West B district | 332 | 13.0 |
| **Stone town** | **901** | **35.3** |
| Wete district | 251 | 9.8 |
| Micheweni district | 195 | 7.6 |
| **Pemba North** | **446** | **17.4** |
| Chakechake district | 219 | 8.6 |
| Mkoani district | 218 | 8.5 |
| **Pemba South** | **437** | **17.1** |

#### Prevalence of Anemia stratified by age group

Overall, almost half of school children and adolescents 5-19 years in Zanzibar were anaemic with a prevalence of 45.7 per cent (Figure 1). Out of these, 24.2%, 21.1% and 0.4% had mild, moderate and severe anaemia respectively. Anaemia was higher (54.6%) among 15-19 years old adolescent girls compared to 38.5% among the boys of the same age group.

**Figure 1.**
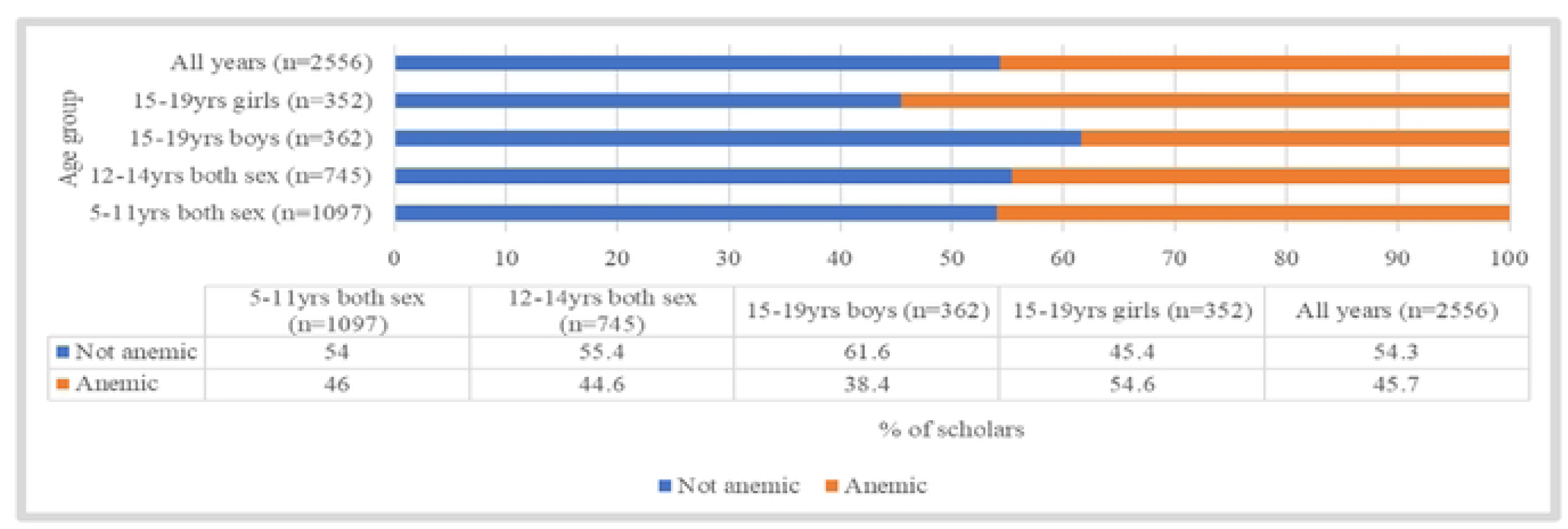
Prevalence of anaemia among children and adolescents aged 5-19 years in Zanzibar.

Figure 2 below describes the level of anaemia among children and adolescents aged 5 to 19 years. Almost 1 in 4 children had mild anaemia followed by moderate (21.1%). The prevalence of severe anaemia was 0.4% with slight variation according to age.

**Figure 2.**
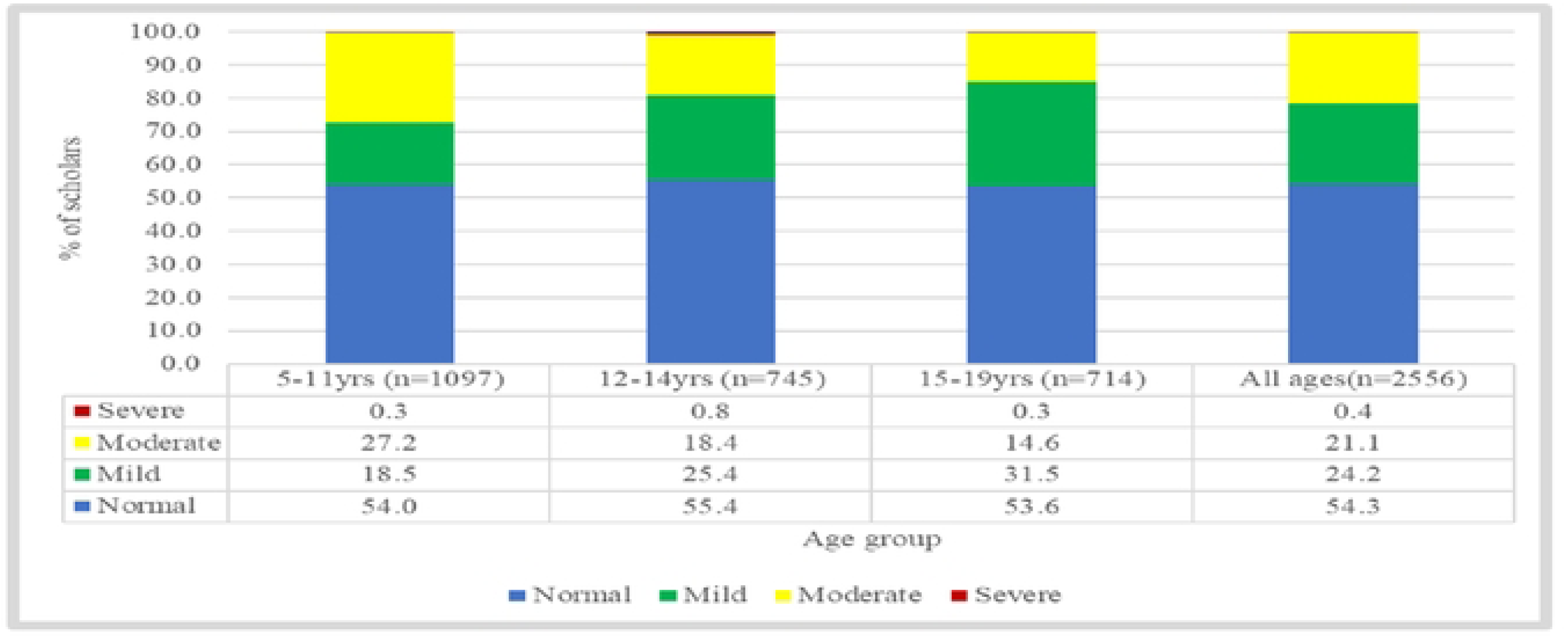
Level of anaemia among children and adolescents 5-19 years in Zanzibar.

#### Prevalence of anaemia stratified by social and demographic characteristics

According to Table 3 below, the prevalence and patterns of anaemia varied across various social and demographic characteristics. The majority of the girls in both age groups were anaemic compared to boys. Overall, 46.0%, 44.6% and 43.6% of school children and adolescents aged 5- 11, 12-14 and 15-19 years respectively were anaemic. The prevalence of anaemia did not vary across settings.

**Table 3:**
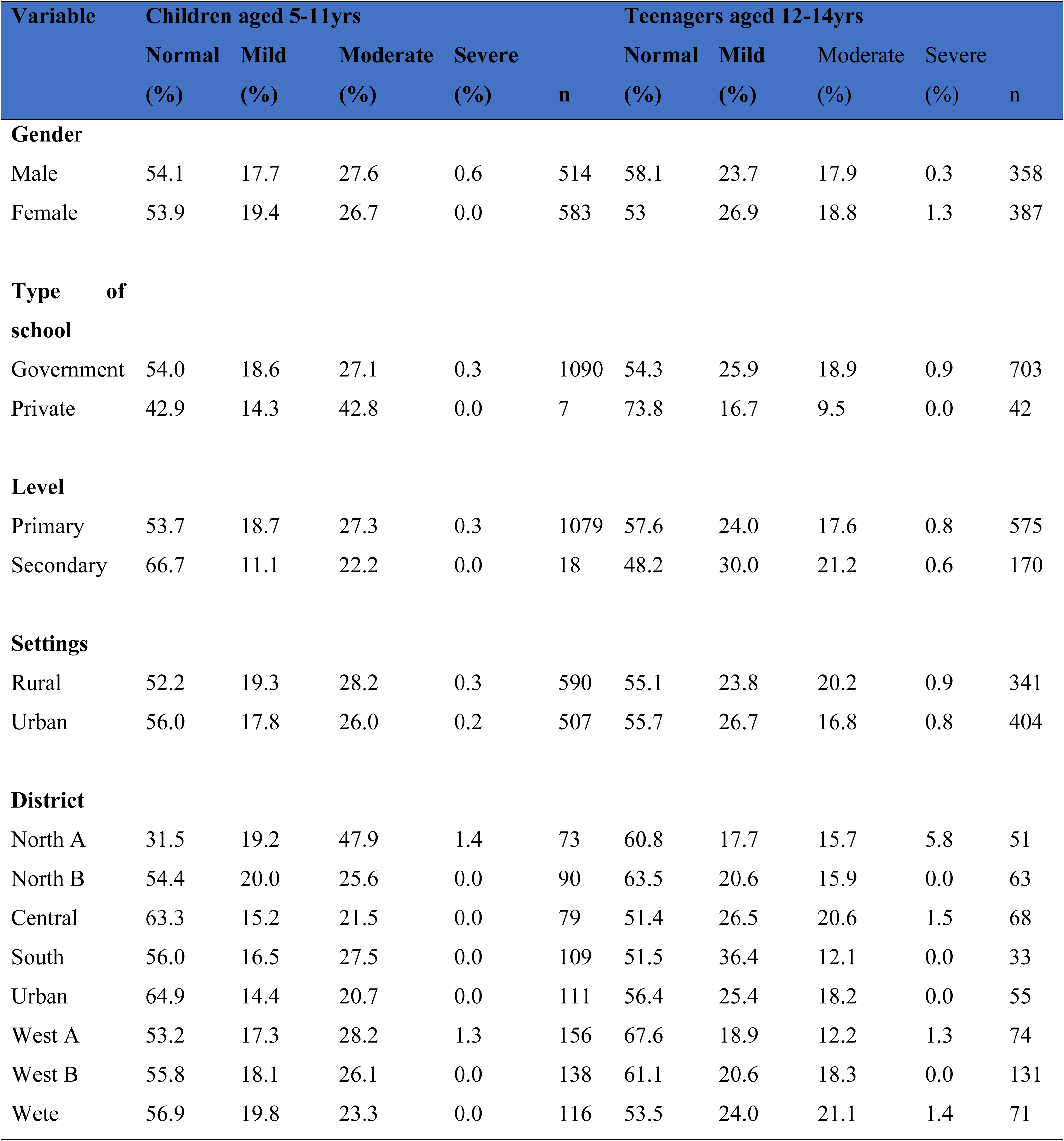
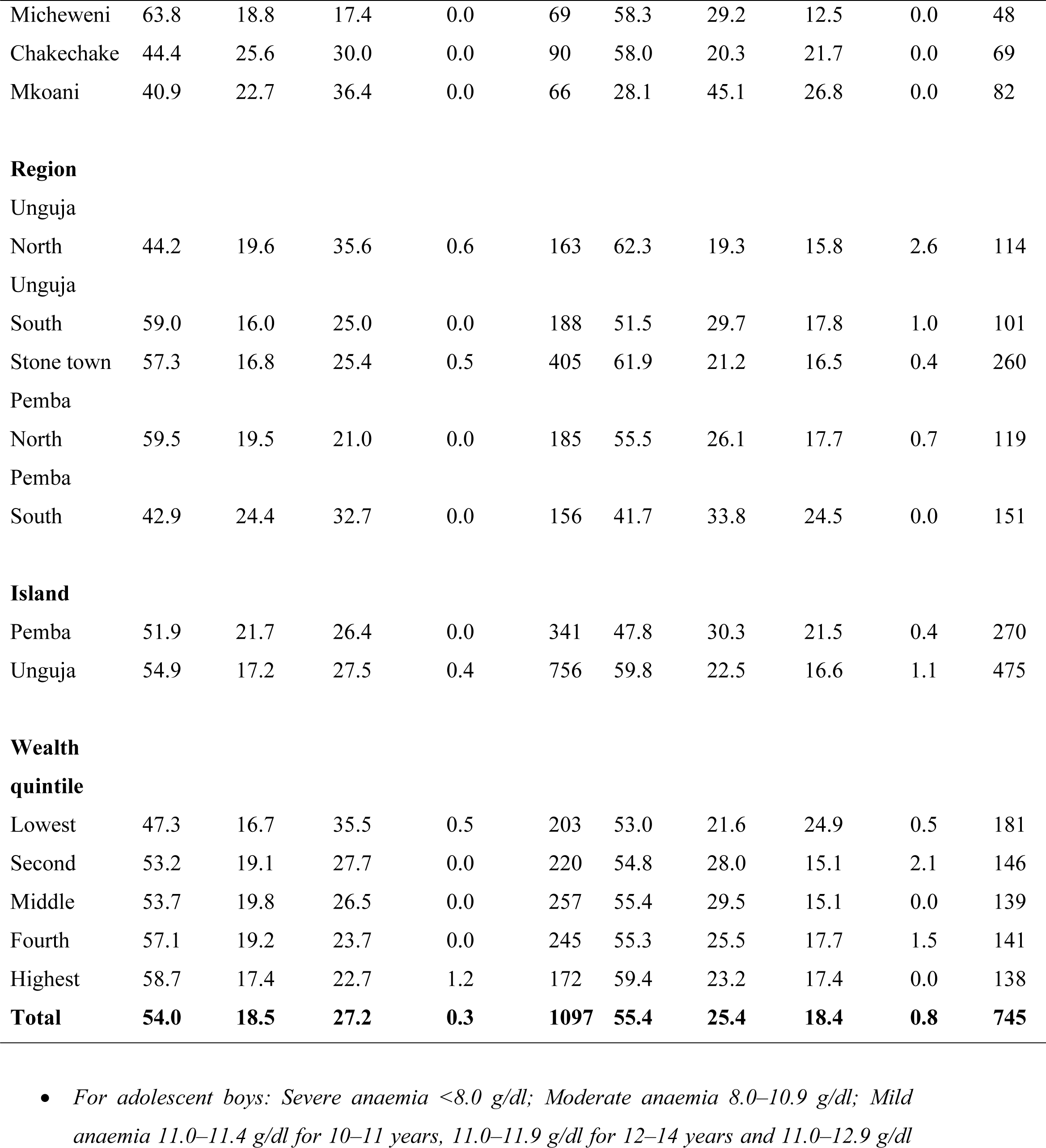
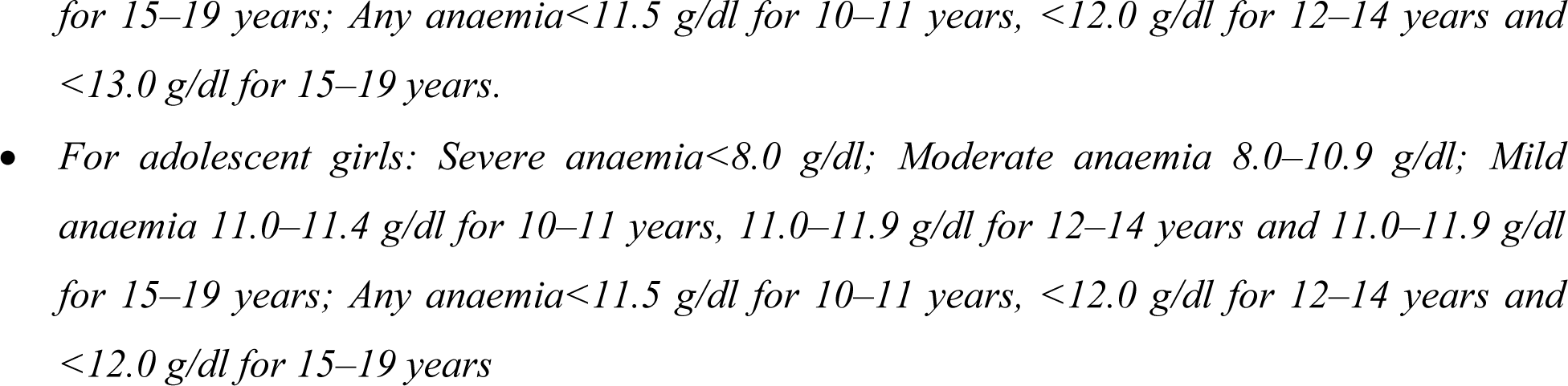
Prevalence of Anaemia among children and school adolescents aged 5–19 years stratified by background characteristics, ZSHNS 2022 (n=2556)

**Table 3:**
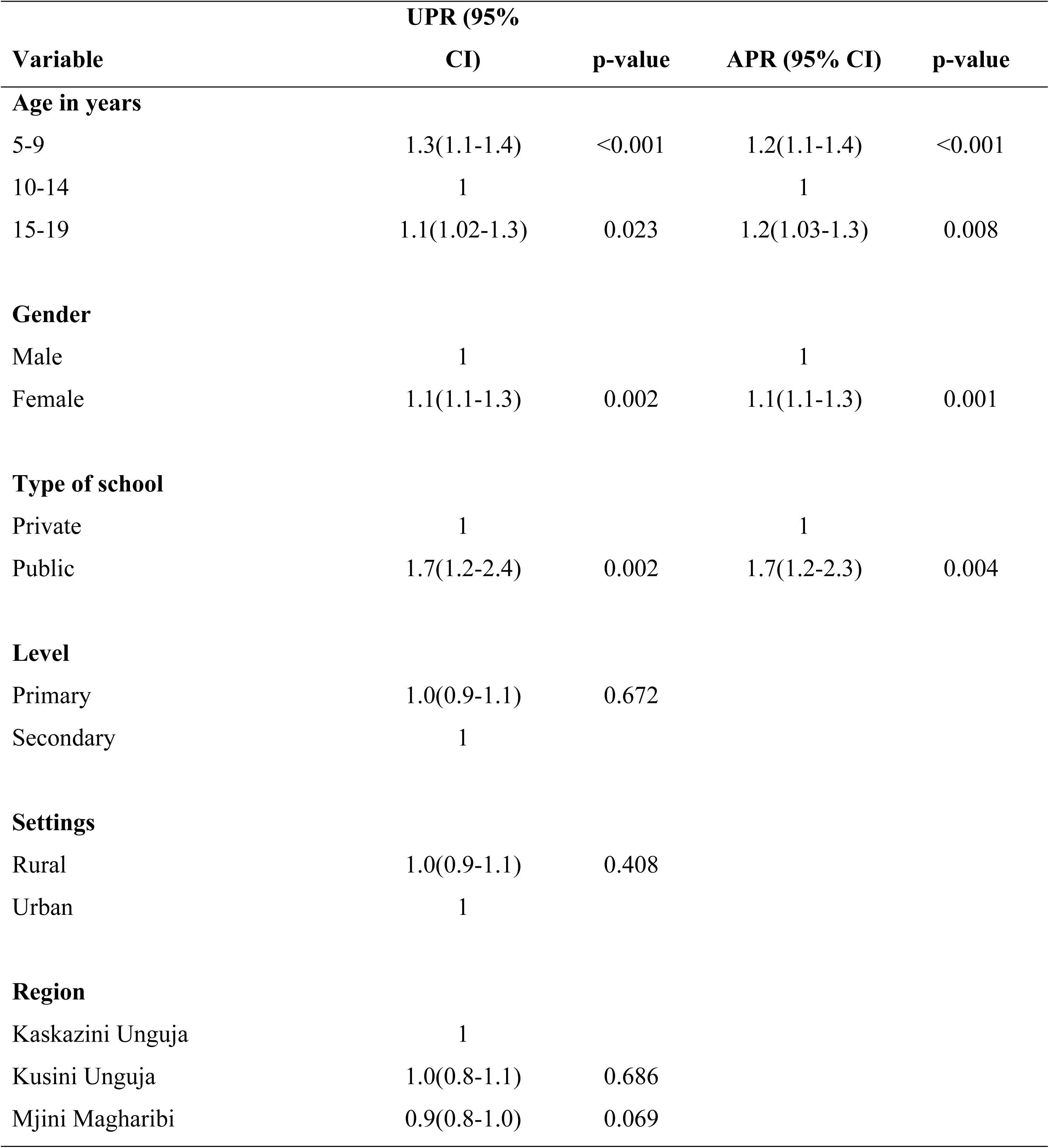
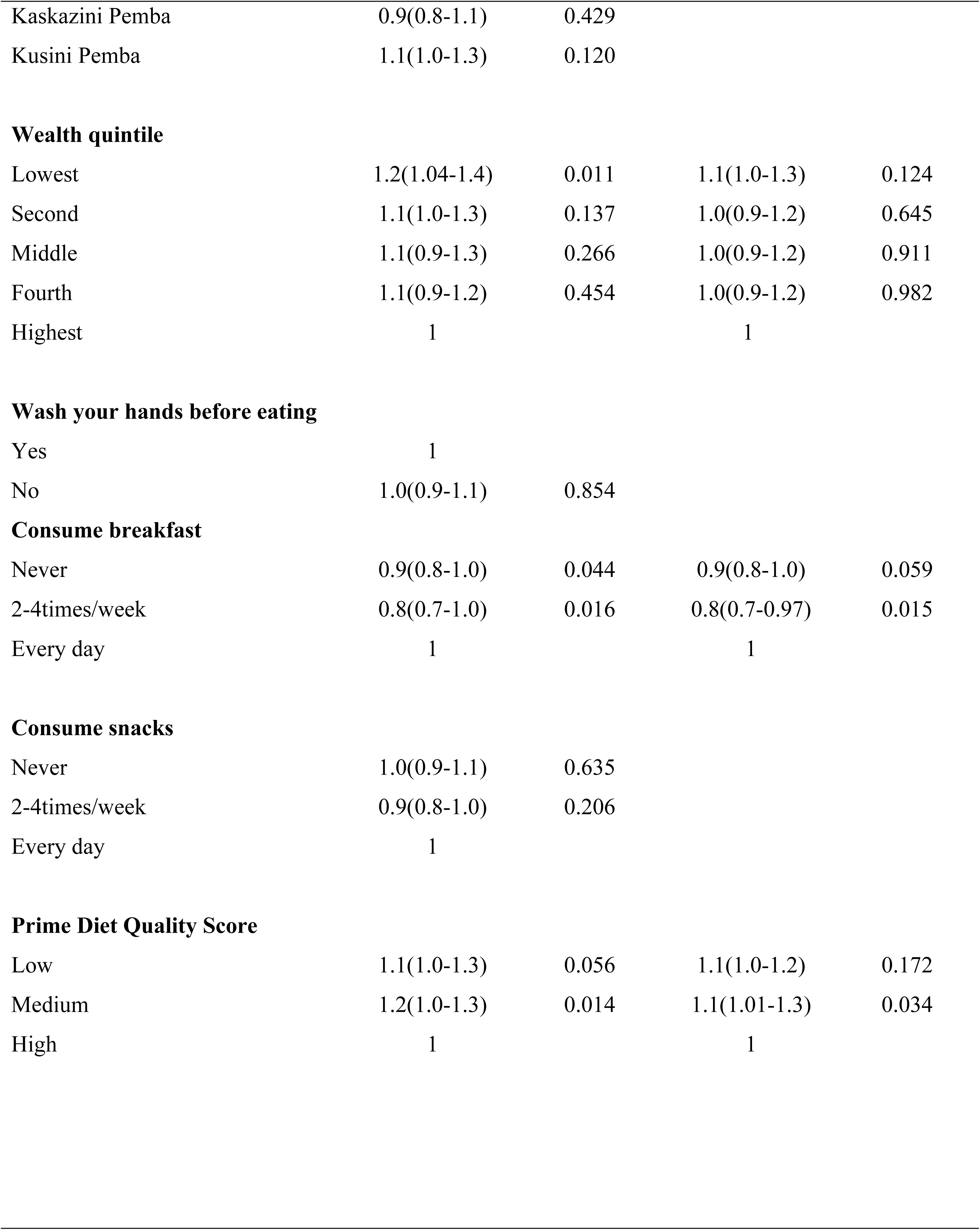
Socio-economic and demographic determinants of anaemia among school children and adolescents in Zanzibar.

| **Variable** | **UPR (95% CI)** | **p-value** | **APR (95% CI)** | **p-value** |
| --- | --- | --- | --- | --- |
| **Age in years** |  |  |  |  |
| 5-9 | 1.3(1.1-1.4) | <0.001 | 1.2(1.1-1.4) | <0.001 |
| 10-14 | 1 |  | 1 |  |
| 15-19 | 1.1(1.02-1.3) | 0.023 | 1.2(1.03-1.3) | 0.008 |
| **Gender** |  |  |  |  |
| Male | 1 |  | 1 |  |
| Female | 1.1(1.1-1.3) | 0.002 | 1.1(1.1-1.3) | 0.001 |
| **Type of school** |  |  |  |  |
| Private | 1 |  | 1 |  |
| Public | 1.7(1.2-2.4) | 0.002 | 1.7(1.2-2.3) | 0.004 |
| **Level** |  |  |  |  |
| Primary | 1.0(0.9-1.1) | 0.672 |  |  |
| Secondary | 1 |  |  |  |
| **Settings** |  |  |  |  |
| Rural | 1.0(0.9-1.1) | 0.408 |  |  |
| Urban | 1 |  |  |  |
| **Region** |  |  |  |  |
| Kaskazini Unguja | 1 |  |  |  |
| Kusini Unguja | 1.0(0.8-1.1) | 0.686 |  |  |
| Mjini Magharibi | 0.9(0.8-1.0) | 0.069 |  |  |
| Kaskazini Pemba | 0.9(0.8-1.1) | 0.429 |  |  |
| Kusini Pemba | 1.1(1.0-1.3) | 0.120 |  |  |
| **Wealth quintile** |  |  |  |  |
| Lowest | 1.2(1.04-1.4) | 0.011 | 1.1(1.0-1.3) | 0.124 |
| Second | 1.1(1.0-1.3) | 0.137 | 1.0(0.9-1.2) | 0.645 |
| Middle | 1.1(0.9-1.3) | 0.266 | 1.0(0.9-1.2) | 0.911 |
| Fourth | 1.1(0.9-1.2) | 0.454 | 1.0(0.9-1.2) | 0.982 |
| Highest | 1 |  | 1 |  |
| **Wash your hands before eating** |  |  |  |  |
| Yes | 1 |  |  |  |
| No | 1.0(0.9-1.1) | 0.854 |  |  |
| **Consume breakfast** |  |  |  |  |
| Never | 0.9(0.8-1.0) | 0.044 | 0.9(0.8-1.0) | 0.059 |
| 2-4times/week | 0.8(0.7-1.0) | 0.016 | 0.8(0.7-0.97) | 0.015 |
| Every day | 1 |  | 1 |  |
| **Consume snacks** |  |  |  |  |
| Never | 1.0(0.9-1.1) | 0.635 |  |  |
| 2-4times/week | 0.9(0.8-1.0) | 0.206 |  |  |
| Every day | 1 |  |  |  |
| **Prime Diet Quality Score** |  |  |  |  |
| Low | 1.1(1.0-1.3) | 0.056 | 1.1(1.0-1.2) | 0.172 |
| Medium | 1.2(1.0-1.3) | 0.014 | 1.1(1.01-1.3) | 0.034 |
| High | 1 |  | 1 |  |

#### Determinants of Anaemia among children and adolescents aged 5 to 19 years

Anemia was 1.2-fold greater (APR: 1.2, 95% CI: 1.1-1.4; p<0.001) if the student was 5-9 years, and as well 1.2-fold higher among 15-19 years students (APR: 1.2, 95% CI: 1.03-1.3; p=0.008). The proportion of anemia was 10% greater among the girls than it was for boys (APR: 1.1, 95% CI: 1.1-1.3; p=0.001). Pupils in public schools had a 1.7 times significantly higher prevalence ratio of having anaemia than their counterparts students from private schools (APR: 1.7, 95% CI: 1.2- 2.3; p=0.004). Pupils who consumed breakfast 2-4 times a week had a significantly decreased prevalence ratio of anaemia (APR: 0.8, 95% CI: 0.7-1.0; p=0.015), while those who never consumed breakfast displayed a non-significant reduced risk of anaemia, with an APR of 0.9 (95% CI: 0.8-1.0; p=0.059). Wealth quintile, school level, regional differences, urban and rural settings, and deworming status, showed no significant difference in the prevalence of anemia.

## DISCUSSION

The Zanzibar National School Health and Nutrition Survey was conducted to fill a knowledge gap where studies on anaemia prevalence among school children appear limited with those in Tanzania Mainland reporting higher prevalence [11,16–17].

The survey found that almost half of the school children and adolescents 5-19 years were anemic.

The implications of these findings are two fold. First, although the prevalence is generally, high, on the positive side, our findings point to a significant reduction in anaemia prevalence among school children and adolescents in Zanzibar. For instance, a study conducted about 26 years ago in Pemba by Stoltzfus et al [18] indicated an overall prevalence of 62.3%. Similarly, a significant reduction of anaemia among children 6–59 months in Zanzibar was reported in a recent study by Said et al [19]Although the age group of 6-59 months in this study is different from the present survey (5-19 years) the findings indicate a significant decrease in anaemia prevalence from 76.1% in 2005 to 65.4% in 2015. This points to a suggestion that efforts to reduce anemia among children in Zanzibar are being productive although the pace is slow. This explains why previous studies continue to recommend the need for designing and strengthening novel and comprehensive interventions to address anaemia both among school children and the general population [18–19].

The second implication of these findings is that the noted prevalence appears to differ from previous studies in Tanzania Mainland and beyond. On the one hand, the prevalence of anaemia among school children in Zanzibar in our study appears to be higher as compared to the findings of some previous studies in Tanzania mainland. For example, a recent cross-sectional study by Mrimi et al [17] conducted among children attending primary schools in Kikwawila and Kiberege wards of South Eastern Tanzania reported an overall prevalence of anemia at 14%. In support, a recent cross-sectional study conducted by Gemechu et al [20] to determine the prevalence of anaemia among school-aged children in Ethiopia reported a prevalence of 24.5%. Another study by Mboera et al [21] among school children in Kilosa documented a prevalence of anaemia of 43.4%. These findings support the premise that the prevalence of anaemia in school-aged children in Zanzibar is higher than in some of the communities in Tanzania Mainland and beyond. This may partly explain why remedial interventions are persistently recommended in previous studies [17–18].

On the other hand, the prevalence of anaemia among school children and adolescents in Zanzibar in this survey appears to be lower as compared to the findings of some previous studies in Zanzibar, Tanzania mainland and beyond. For instance, anaemia studies conducted in Tanzania between 2015 and 2023 with a focus on school children and under-five children have persistently reported higher prevalences [21–23]. Previous studies of anaemia in children in Zanzibar [18–19], Mwanza [11] and Arusha [23] for example have reported a prevalence ranging from 62.3% to 84.6%. In support, a recent multicountry study by Partap et al [22] indicated a prevalence of anaemia of 58.3% in Tanzania which was almost 5 times of Ethiopia and more than 2 times of Sudan. This indicates that the prevalence of anaemia among school children may be less than that reported in some studies within and beyond Tanzania. However, this does not underscore the value of designing novel and integrated interventions to reduce the burden of anaemia among children in Zanzibar.

Other important findings of this survey are the determinants of anaemia among school children and adolescents in Zanzibar. This survey indicated that anaemia in school children and adolescents is strongly associated with gender, age, school ownership, meal frequency and family income. Children aged 5-9 years, female children, children in public schools, children who consume breakfast 2-4 times a week, and those in the lowest wealthy quantiles were more likely to be anaemic but this was found not significant when adjusted for other factors. Again, the implications of these findings are two fold. First, the determinants of anaemia among school children and adolescents in this survey appear to be slightly different from those in previous studies within Zanzibar, Tanzania mainland and beyond. For example, a 26-year-old study in Zanzibar [18] linked anaemia to gender with boys, older (above 11 years) and younger children (below 7 years), hookworm infestations and stunted children being likely to be anaemic. Likewise, another study in Zanzibar [19] linked anaemia to improper stool disposal practices, poor dietary diversity, low age and underweight. These findings are different from those reported in the current survey where girls, children aged 5-9 years, poor uptake of breakfast and low income were linked to anaemia. The variations in determinants of anaemia in these studies may be partly explained by the primary age of focus e.g. 6-59 months [19] and the distribution of anemia [18]. Nevertheless, the variations of determinants of anaemia in Zanzibar appeared to extend to Tanzania’s Mainland and beyond. With most focusing on under-five children, studies in Tanzania Mainland have linked anaemia to low birth weight, not consuming meat and vegetables, drinking milk and tea [23]; Malaria and micronutrient deficiency [17, 24], HIV infection [24] and hookworm infections [25]. Again, the age of focus in most of these studies (6-59 months) may partly explain the variations.

Second, some of the findings of this survey are slightly similar to the findings of previous studies with minimal variations. For example, a study by Lwambo [25] among primary school students (aged 7-18 years) in Mwanza noted a high prevalence of anemia, which decreased with an increase in age and gender, with boys having a lower likelihood of anemia compared to girls. The lower likelihood of anemia among boys compared with girls is somewhat similar to the findings of this survey, which reported that girls were more likely to be anemic as compared to boys. This suggests that studies that focus on school children with almost similar age groups to our study may report similar determinants of anaemia in other parts of Tanzania. The high prevelance of anemia among gilrs call for more targeted interventions such as iron and foloc acid supplementation among adolescents girls as recommended by the World Health Organization [16, 17].

## Limitations

This study is not without limitations. Although the findings address the research gap in Zanzibar and may be useful in policy formulation, the study did not examine the contributors of anaemia on the caregiver’s and community’s side. Further studies that tap into the insights of community members on the determinants of anaemia are recommended for generating a comprehensive understanding of the persistent contributors of anaemia among children in Zanzibar.

## Conclusion

Despite massive improvements as compared to previous studies, the findings indicate a persistently higher prevalence of anaemia among school children and adolescents in Zanzibar. The prevalence of anaemia among school children in Zanzibar is higher than in some communities which points to the need for novel strategies for reducing its burden. The preponderance of anaemia among adolescent girls and those from public schools provides a unique opportunity for targeting such populations with anaemia prevention and control interventions, and schools are uniquely positioned to provide such interventions.

## Data Availability

Data will be shared with the authorization of the author

## Acknowledgement

The Ministry of Health (MoH) Zanzibar would like to express its gratitude and appreciation to all individuals, institutions, and partners who contributed to the implementation of this School Health and Nutrition Survey (SHNS) conducted in Zanzibar. In particular, the MoH acknowledges the contribution of partners from governmental agencies, the Office of the Chief Government Statistician (OCGS), the Ministry of Education and Vocational Training (MoEVT) together with the Tanzania Food and Nutrition Centre for their active participation and technical inputs toward this survey.

Special thanks go to the United Nations Children’s Fund (UNICEF) for their technical and financial support, and for seeing the requirement to adopt a similar study on Tanzania Mainland. The MoH would also like to thank all individual consultants, oversight organisations and partners for their roles in organising and preparing questionnaires, training of field staff, co-ordination of fieldwork, and support in developing this report, who include individuals from the Muhimbili University of Health and Allied Sciences (MUHAS), the National Institute for Medical Research (NIMR), and the Nelson Mandela University.

The MoH would also like to thank all the field supervisors, who oversaw the data collection exercise, and field data collectors who interviewed students. Moreover, the MoH would like to extend its appreciation to Data Entry Clerks, Regional and District teams (e.g., Malaria Focal Persons, laboratory technologists/technicians, School Health Programme Co-ordinators, Nutrition Officers, and drivers) for their participation during the survey. Furthermore, the MoH would like to thank all the students and school teachers for their readiness to participate in the survey. The MoH would also like to acknowledge the following teams of experts involved in the report preparation and finalization.

Data analysis, Report writing and finalisation Team: Dr. Geofrey Mchau, Sauli Epimark, Shemsa Msellem, Hawa Msola, Fatma Abdallar, Dr Anna Mosses, Dr David Solomon and Asha Samini, Fatma Said, Adam Haji Ame, Heavenlight Ayubu, Stanlaus Erick Mollel, Salum Kassim Ally, Fahima Mohamed Issa, Sabina Raphael Daima, Kombo Mdachi Kombo, Hadija Hamisi Hamadi, Ahmad Hamza Mohamed, Aisha Mohamed Said, Asha Mahafodhi, and Prof. Esther Ngadaya.

